# PixelPrint: Three-dimensional printing of realistic patient-specific lung phantoms for validation of computed tomography post-processing and inference algorithms

**DOI:** 10.1101/2022.05.06.22274739

**Authors:** Nadav Shapira, Kevin Donovan, Kai Mei, Michael Geagan, Leonid Roshkovan, Grace J. Gang, Mohammed Abed, Nathaniel Linna, Coulter Cranston, Cathal O’Leary, Ali Dhanaliwala, Despina Kontos, Harold I. Litt, J. Webster Stayman, Russell T. Shinohara, Peter B. Noël

## Abstract

**Background:** Radiomics and other modern clinical decision-support algorithms are emerging as the next frontier for diagnostic and prognostic medical imaging. However, heterogeneities in image characteristics due to variations in imaging systems and protocols hamper the advancement of reproducible feature extraction pipelines. There is a growing need for realistic patient-based phantoms that accurately mimic human anatomy and disease manifestations to provide consistent ground-truth targets when comparing different feature extraction or image cohort normalization techniques.

**Materials and Methods:** PixelPrint was developed for 3D-printing lifelike lung phantoms for computed tomography (CT) by directly translating clinical images into printer instructions that control the density on a voxel-by-voxel basis. CT datasets of three COVID-19 pneumonia patients served as input for 3D-printing lung phantoms. Five radiologists rated patient and phantom images for imaging characteristics and diagnostic confidence in a blinded reader study. Linear mixed models were utilized to evaluate effect sizes of evaluating phantom as opposed to patient images. Finally, PixelPrint’s reproducibility was evaluated by producing four phantoms from the same clinical images.

**Results:** Estimated mean differences between patient and phantom images were small (0.03-0.29, using a 1-5 scale). Effect size assessment with respect to rating variabilities revealed that the effect of having a phantom in the image is within one-third of the inter- and intra-reader variabilities. PixelPrint’s production reproducibility tests showed high correspondence among four phantoms produced using the same patient images, with higher similarity scores between high-dose scans of the different phantoms than those measured between clinical-dose scans of a single phantom.

**Conclusions:** We demonstrated PixelPrint’s ability to produce lifelike 3D-printed CT lung phantoms reliably. These can provide ground-truth targets for validating the generalizability of inference-based decision-support algorithms between different health centers and imaging protocols, as well as for optimizing scan protocols with realistic patient-based phantoms.

## INTRODUCTION

Quantitative imaging is receiving increased interest and acknowledgment from clinicians and healthcare providers as a supporting tool for data-driven, patient-specific clinical decision making^1–3^. Driven by a pursuit for precision medicine, developments focus on identifying biomarkers that are invisible to the naked eye but can be used for evidence-based inference for clinical decision support or to establish reliable correlations between image features and clinical outcomes, prognosis assessments, and treatment response predictions^4,5^. However, variability in image acquisition and reconstruction techniques introduce heterogeneity in image characteristics and features that are independent of the underlying biology and pathophysiology^6^. Modern medical imaging modalities, such as computed tomography (CT), magnetic resonance imaging (MRI), and positron emission tomography (PET), allow a wide variety of imaging parameters that are, in general, lacking standardization between different health centers and different scanner models. While these differences typically have little clinical impacts for routine radiological interpretation, they introduce biases when analyzed numerically to extract meaningful data^6^. This hampers advancement of reproducible feature extraction pipelines, a critical pre-requisite for clinical translation^7^.

Despite ongoing efforts to account for factors originating from the recognized lack of imaging standardization, the problem of biases and variability persists. Experimental validation of image cohort normalization methods, such as ComBat^8–10^, is currently limited due to an inability to repeat patient scans on multiple scanners or with multiple imaging protocols given logistical and risk-related considerations, e.g., the risks of ionizing radiation in CT and PET. There is therefore a growing need for realistic patient-based volumetric phantoms that can accurately mimic human anatomy and disease manifestations to provide consistent imaging ground-truth targets when comparing post-processing image cohort normalization and feature extraction techniques.

Anthropomorphic phantoms are fundamental tools for developing, optimizing, and evaluating hardware and software advances in medical imaging research and clinical practice. Such phantoms are typically manufactured by machining, casting, or molding homogenous materials that mimic tissue properties relevant for the specific imaging modality, e.g., x-ray attenuation coefficients for CT^11^. Realistic patient-based phantoms have additional advantages for clinical and development tasks, such as imaging protocol optimization, and provide ground-truth targets for denoising or artifact correction AI algorithms. Despite a wide range of commercially available phantoms, there is a lack of patient-based phantoms capable of reliably representing the quantitative imaging characteristics and textures found in clinical patient images. The academic and clinical radiology communities would greatly benefit from rapid, versatile, lifelike, as well as inexpensive phantom manufacturing processes, compared to commercial solutions currently available.

Throughout the last decade, three-dimensional (3D)-printing of phantoms that represent the x-ray attenuations and textures of various tissues, anatomies, and disease has been widely explored. These studies focused on several developmental aspects, including 3D-printing of accurate attenuation profiles^12–15^, manufacturing anatomically-correct organ models^16–20^, and generation of realistic tissue textures^21–23^. Novel 3D-printing techniques, mainly using fused deposition modeling (FDM), have been proposed to generate variable material densities that mimic the imaging features observed in clinical CT images. These methods^24–26^ include utilization of different infill printing patterns^27^, variable voxel-dependent extrusion rates^14,15^, or interlacing two different materials with dual-extrusion printers^28^.

Generation of 3D-printed anthropomorphic phantoms from clinical CT images typically involves^19,29–32^: (i) automated or manual segmentation of specific tissues or organs, e.g., an entire lung or identified findings, (ii) conversion of the segmented volumes into triangulated surface geometry models, such as standard triangle/tessellation language (STL), and (iii) utilization of printer-specific slicing software to generate instructions (e.g., G-code) that determine relevant 3D-printing parameters, such as extrusion rate, printing speed, infill ratios, etc. While phantoms produced this way may approximate clinical imaging characteristics, they still have shortcomings. Most importantly, due to segmentation of regions followed by conversion to surface models, abrupt and unrealistic transitions between homogenous regions of different densities are created within the printed products, and spatial resolution and textural information are compromised.

In this work we evaluate a promising alternative called PixelPrint that we recently developed to overcome the limitations described above. PixelPrint directly translates DICOM image data into printer instructions that continuously control the printed material density by varying the printer speed on a voxel-by-voxel basis, while maintaining a constant filament extrusion rate^33^. We report on reader studies conducted to assess the correspondence between imaging characteristics of three 3D-printed COVID-19 pneumonia lung phantoms with those of the original patient images used to produce these phantoms. We also report quantitative comparisons between four 3D-prints of the same patient for production reproducibility assessments.

## METHODS

Three patient cases were selected from the Hospital of the University of Pennsylvania PACS by a thoracic radiologist (LR, four years of experience) under an IRB approved protocol. Patients were selected based on the assessed COVID-19 severity level (mild, moderate, severe), patient habitus, and absence of significant metal artifacts. For each patient, clinical DICOM images reconstructed with a sharp kernel (Table 1) were converted into 3D-printer instructions using PixelPrint software. A complete technical background of the PixelPrint algorithm, pipeline, and quantitative evaluation is available in our previous publication^33^. A primary advancement of PixelPrint presented in this study is the 3D-printing of phantoms based on volumetric patient data (Figure 1). All phantoms presented in this work were printed using 1.75 mm diameter Polylactic Acid (PLA) filament (MakeShaper, Keene Village Plastics, Cleveland, OH, USA) on a Lulzbot TAZ 6 fused-filament 3D-printer (Fargo Additive Manufacturing Equipment 3D, LLC Fargo, ND, USA) with a 0.25 mm brass nozzle. Phantoms were printed with a constant extrusion rate of 0.6 mm^3^/sec and a layer height of 0.2 mm. Printing speeds varied from 3 to 30 mm/s, with acceleration and jerk (threshold velocity for applying acceleration) settings of 500 mm/sec^2^ and 8 mm/sec, respectively, producing line widths from 0.1 to 1.0 mm.

**Table 1:**
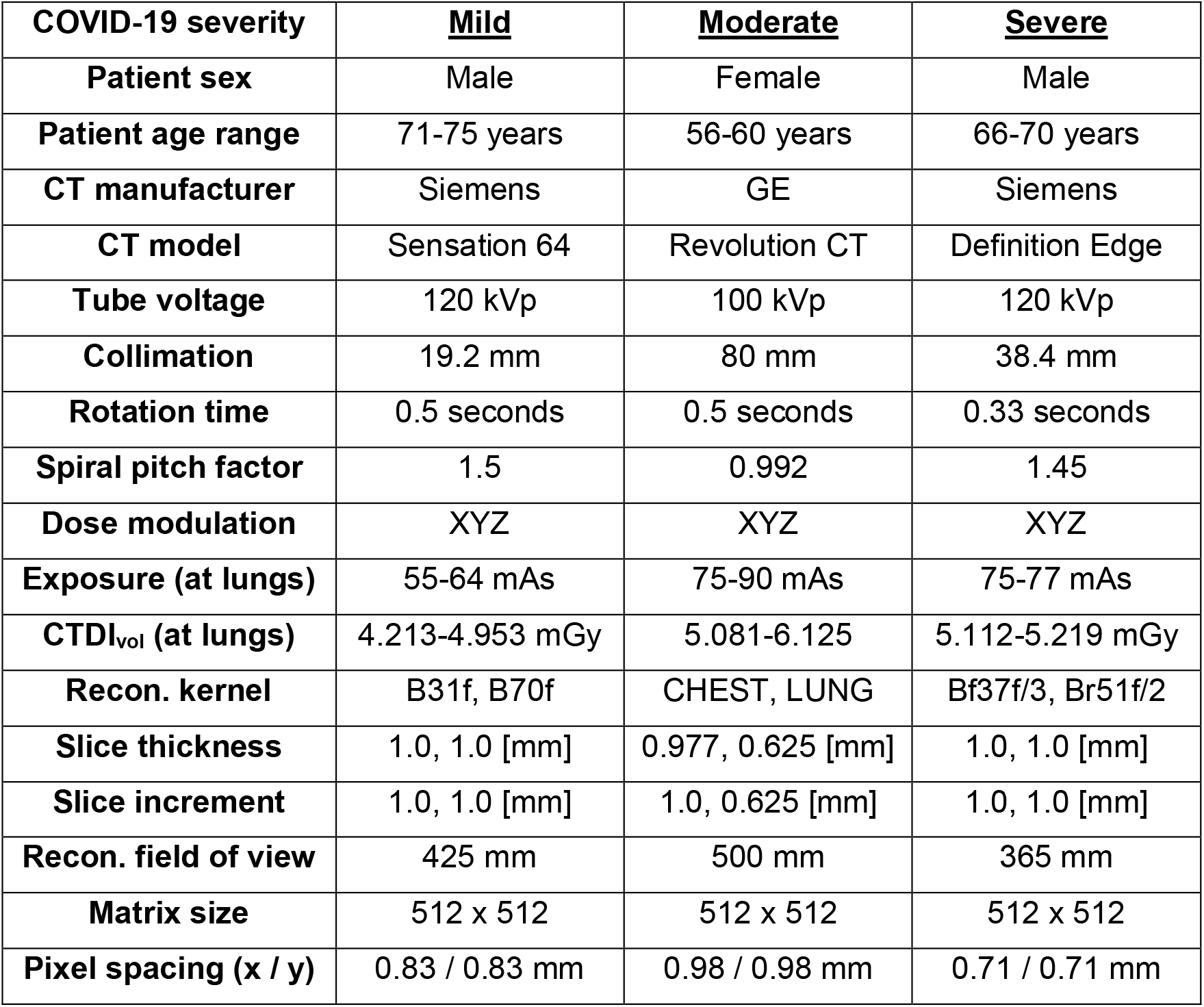
Patient information together with the scan and reconstruction parameters that were used to generate the original diagnostic CT images and the images of the three corresponding 3D-printed phantoms.

**Figure 1:**
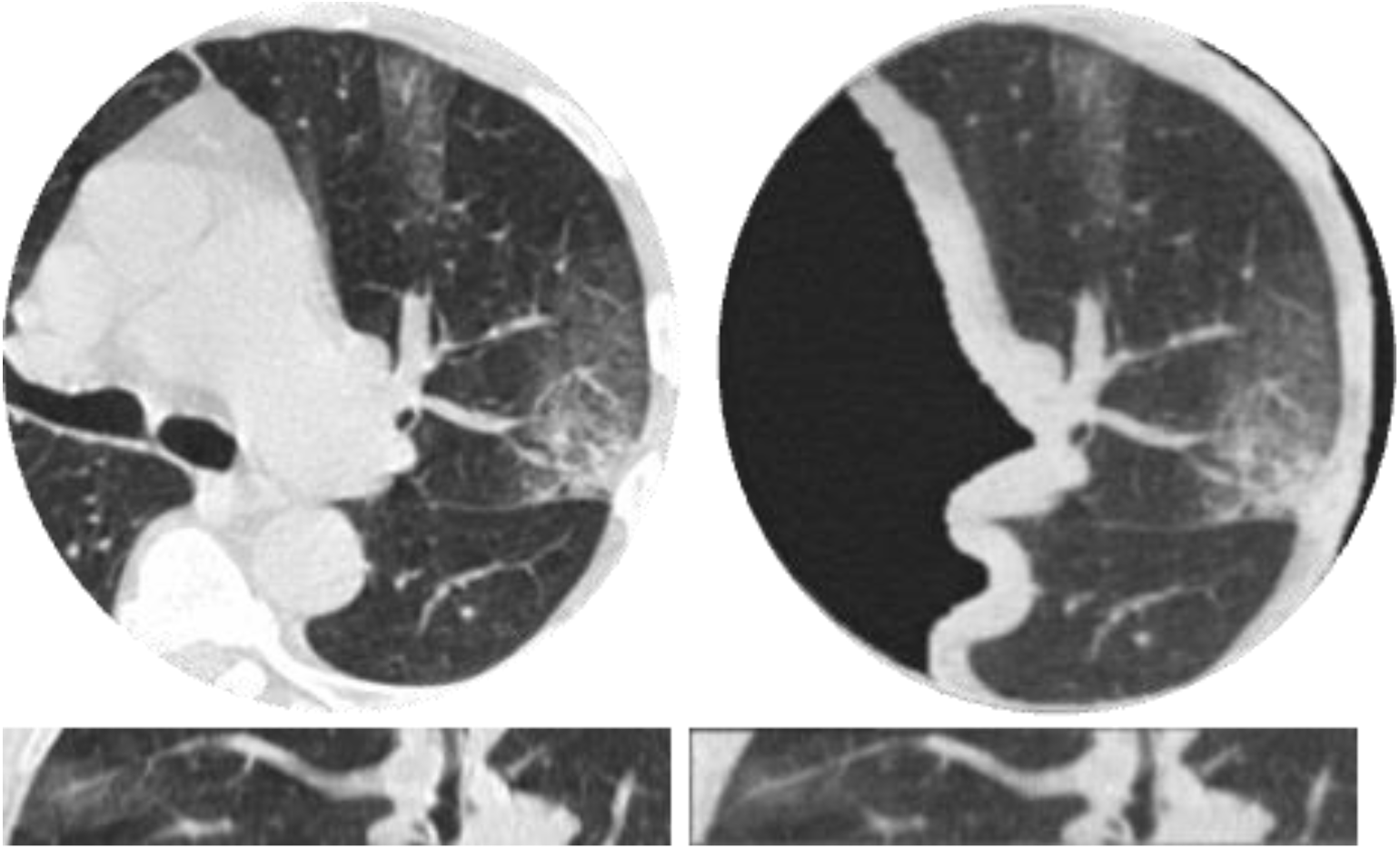
Comparisons between clinical CT lung images of a mild COVID-19 patient (left) and images of a corresponding 30 mm thick 3D-printed volumetric phantom (right), acquired with the same CT scanner and imaging parameters. Presented in two orthogonal views: axial (top), sagittal (bottom). Window level/width are - 500/1400 HU.

Each phantom was scanned on the same scanner using the same acquisition and reconstruction settings as the input patient scan (Table 1). The phantoms were placed within the 20 cm bore of a 300 × 400 mm^2^ phantom (Gammex MECT, Sun Nuclear, Melbourne, FL, USA) to mimic attenuation profiles of a medium sized patient. A preprocessing pipeline was developed for preparing images for a reader study using the following steps. First, lung segmentations obtained using a pretrained AI^34^ from each of the original patient scans were dilated by eight pixels in every direction and manually positioned on the 3D-printed phantom image volumes. Next, an image registration algorithm (Simple-ITK^35^) was applied to accurately align phantom images with their corresponding patient images and a circular binary mask of 18 cm diameter was applied to both the segmented phantom and their corresponding patient images to hide their surroundings (patient anatomy or MECT phantom). Finally, images from both the phantoms and the corresponding patient images were randomized separately each reader evaluation.

The reader study consisted of two parts. In the first part, radiologists were asked to review 120 randomized slices from patient and phantom scans, reconstructed with either a sharp or smooth kernel, and answer four questions regarding whether the presented slice had realistic imaging, contrast, noise, and resolution characteristics of a diagnostic quality CT lung scan. In the second part, radiologists were asked to review 90 randomized slices from patient and phantom scans, all reconstructed with a smooth kernel, and for each slice rate the severity of COVID-19 consolidations (none, mild, moderate, severe) and whether there are sufficient details (e.g., resolution, contrast-to-noise ratios) for a confident COVID diagnosis. To simplify the analysis of the reader study, a higher rating indicates a better review score for all questions except for the COVID-19 severity question. A dedicated user interface was implemented to simplify the review process and to record the radiologists’ replies. Importantly, the participating radiologists were told that they were taking part in a “CT lung image evaluation study” and were completely unaware of the fact that the reviewed datasets included phantom images, which is why this study can be considered a “completely blinded” reader study.

Statistical analysis was performed to assess the mean difference in responses between patient and phantom images with the aid of linear mixed models. For this, each question was modeled (separately) using the following equation:

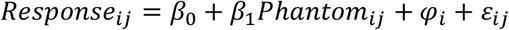

where *i* denotes the reader and *j* denotes the image, *β*_0_ and *β*_1_ denote the mean response across readers for patient scans and difference in mean response between patient and phantom across readers, respectively. The model allows estimation of the mean rating difference between phantom and patient images, while controlling for potential differences between readers in their responses through *φ*_*i*_ and *ε*_*ij*_. *φ*_*i*_, which represents reader-level differences in mean response for a given question, and *ε*_*ij*_, which represents the remaining model errors, are assumed to be independent across scans and readers with equal variance and zero mean, as well as normally distributed.

Along with statistical significance, which was assessed through standard hypothesis testing, a measure of “clinical significance” is important to quantify the estimated difference between the two set of images, i.e., phantom vs. patient, with respect to different measures of variance. This is because while differences may be “statistically” significant based on the resulting p-values, at the same time they may be clinically insignificant in terms of their magnitude relative to inter- and intra-observer variabilities. Moreover, if sample sizes are large, arbitrarily small differences will often be statistically significant^36^. Thus, assessments of effect sizes are critical to fully assess the mean difference^37^. In the two-sample context, Cohen’s *d* is a commonly used measure of effect size^38^. However, in the context of clustered data, where in this case readers are the clusters, a different estimate for the pooled standard deviation is needed. An alternative for this context, proposed in Westfall *et al*., is 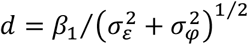 where 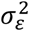 denotes the variance in the error terms (within-reader variance) and 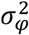 denotes the between-reader variance^39^. Another similar effect size measure is the ratio between the mean difference and within-reader variability, given by *d*′ = |*β*_1_|/*σ*_*ε*_. Both effect size calculations were assessed here as part of our analysis, together with R^2^ calculations to measure the proportion of response variation that is associated with the scanned object type (patient vs. phantom).

Finally, to assess the robustness and reproducibility of PixelPrint’s phantom production process, three additional phantoms were 3D-printed based on the moderate COVID-19 patient images. The four theoretically equivalent phantoms were scanned on a dual-energy CT scanner (IQon, Philips Healthcare, Cleveland, OH, USA) using an axial protocol at 120 kVp and 0.75 seconds rotation time, both at clinical dose exposure levels (6 mGy CTDI_vol_) and at high dose exposure levels (18 mGy CTDI_vol_), and reconstructed with a smooth kernel and a 250 mm field of view at 1.0 mm slice thicknesses. Correspondence between the four phantoms was evaluated with the structural similarity index measure (SSIM).

## RESULTS

To visualize the data, frequency of reader ratings and mean response values are provided in Figures 2 and 3. Figure 2 provides the counts of each response score as values between “1” and “5”, where a higher rating indicates a better score, across all questions and separated between readers. The figure reveals similar counts between the patient and phantom images, with a response of “4” being most common in both cases for both scan types. Figure 3 presents calculated mean ± one standard deviation (SD) response values for each reader and question, separated by the patient COVID-19 severity. Visually, the patient scans have a higher mean response across the different severity levels, however, these differences are small in all cases (<0.5), and are mainly driven by the responses of the first reader.

**Figure 2:**
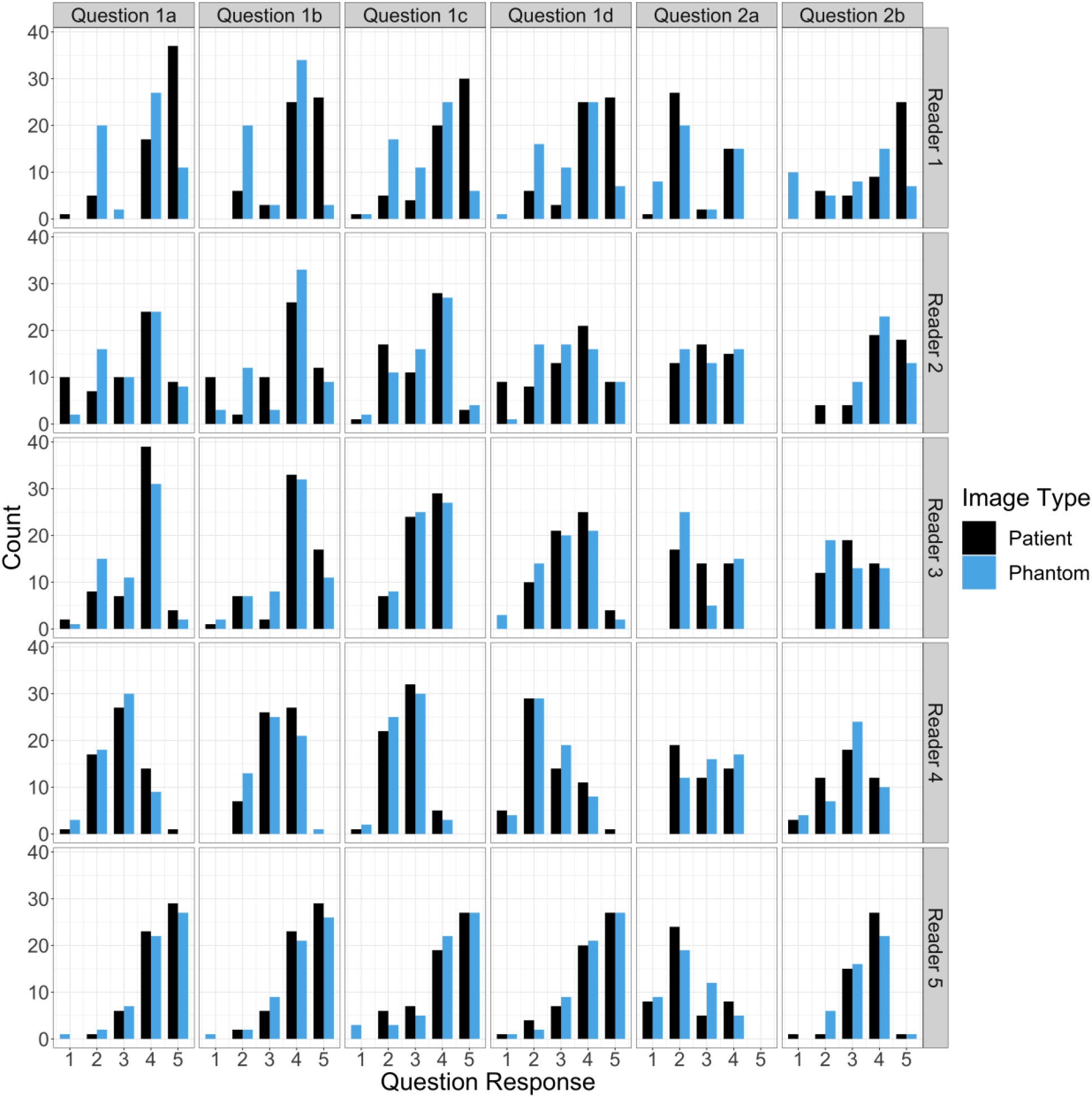
Counts of responses for phantom and patient images by reader (rows) and question (columns): (1a-d) Imaging, contrast, noise, and resolution characteristics; (2a) COVID-19 severity; and (2b) diagnostic confidence. Except for the COVID-19 severity question, higher ratings indicate better review scores. Overall, the count frequencies portray a high correspondence between phantom and patient images.

**Figure 3:**
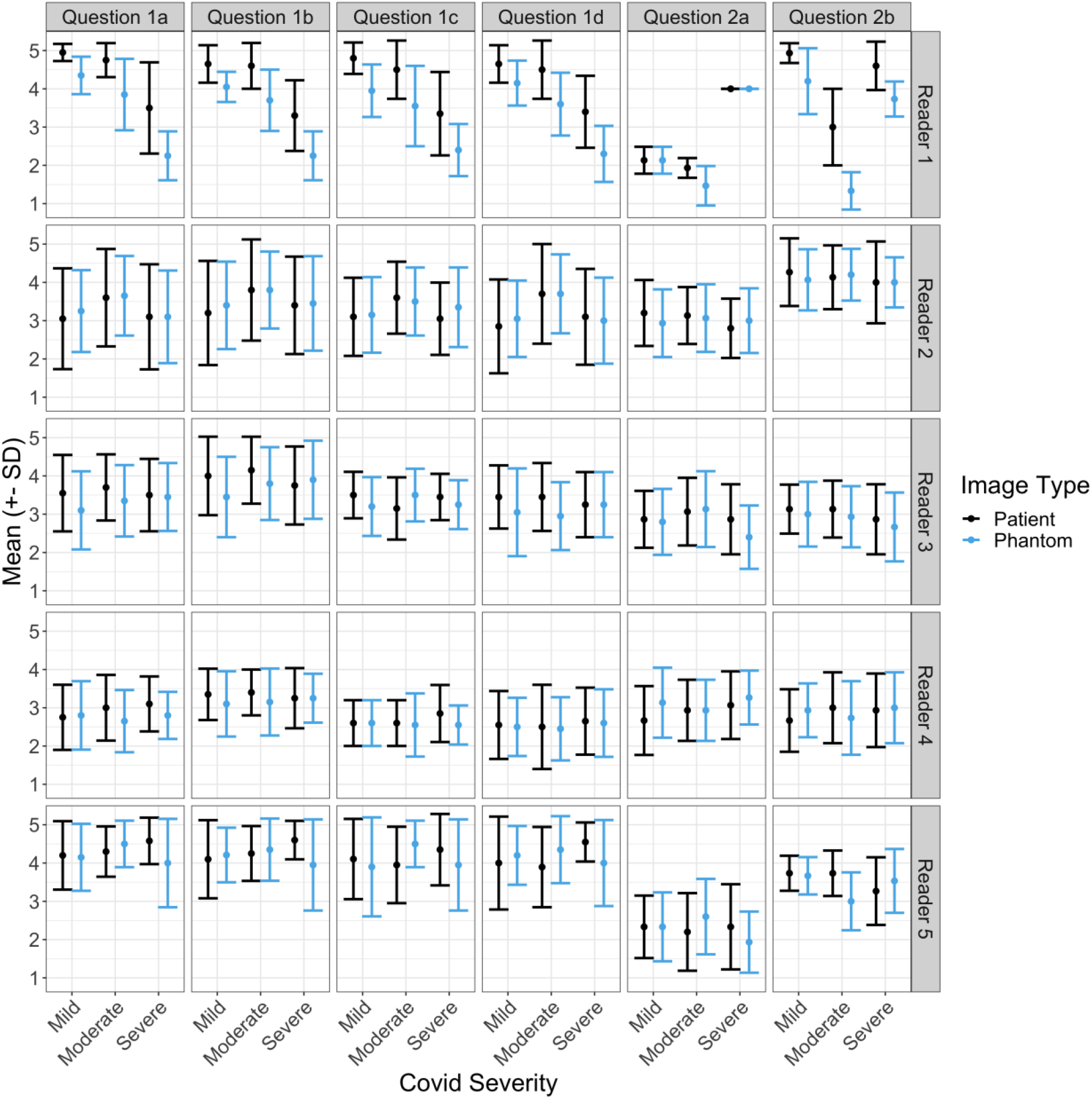
Mean ± standard deviations (SD) of responses for different COVID-19 severity levels on phantom and patient images by reader (rows) and question (columns): (1a-d) Imaging, contrast, noise, and resolution characteristics; (2a) COVID-19 severity; and (2b) diagnostic confidence.

Figure 4 presents differences in reader ratings between phantom slices that have corresponding (paired) patient slices, i.e., differences in rating between a phantom slice and its matching patient slice: same reader, COVID-19 severity, slice location, and convolution kernel (sharp/smooth), together with Gaussian fits to the data. In general, the data indicates rating differences that are centered between -0.04 and 0.38, implying that on average differences in reader ratings between phantom and patient images are much smaller than a single rating point. Modeling results for the six questions that compose both parts of the reader study are provided in Table 2. Each row in the table reports the mean rating (*β*_0_), rating difference between patient and phantom images (*β*_1_), and R-squared values that were obtained for each question separately. Within a question, for a given parameter the estimate, 95% CI, and p-value are provided. Since the rating scores are categorical, p-values for this parameter are not included. In all cases, while the estimated mean differences between patient and phantom were statistically significant (p<0.005), these differences were very small in magnitude, ranging from 0.03 to 0.29. The magnitude of the difference was also evaluated using R^2^ measures, resulting in low values for all questions, with a maximum of 0.02 maximum, indicating that a low proportion of response variation is associated with replacing a patient image with a phantom image.

**Figure 4:**
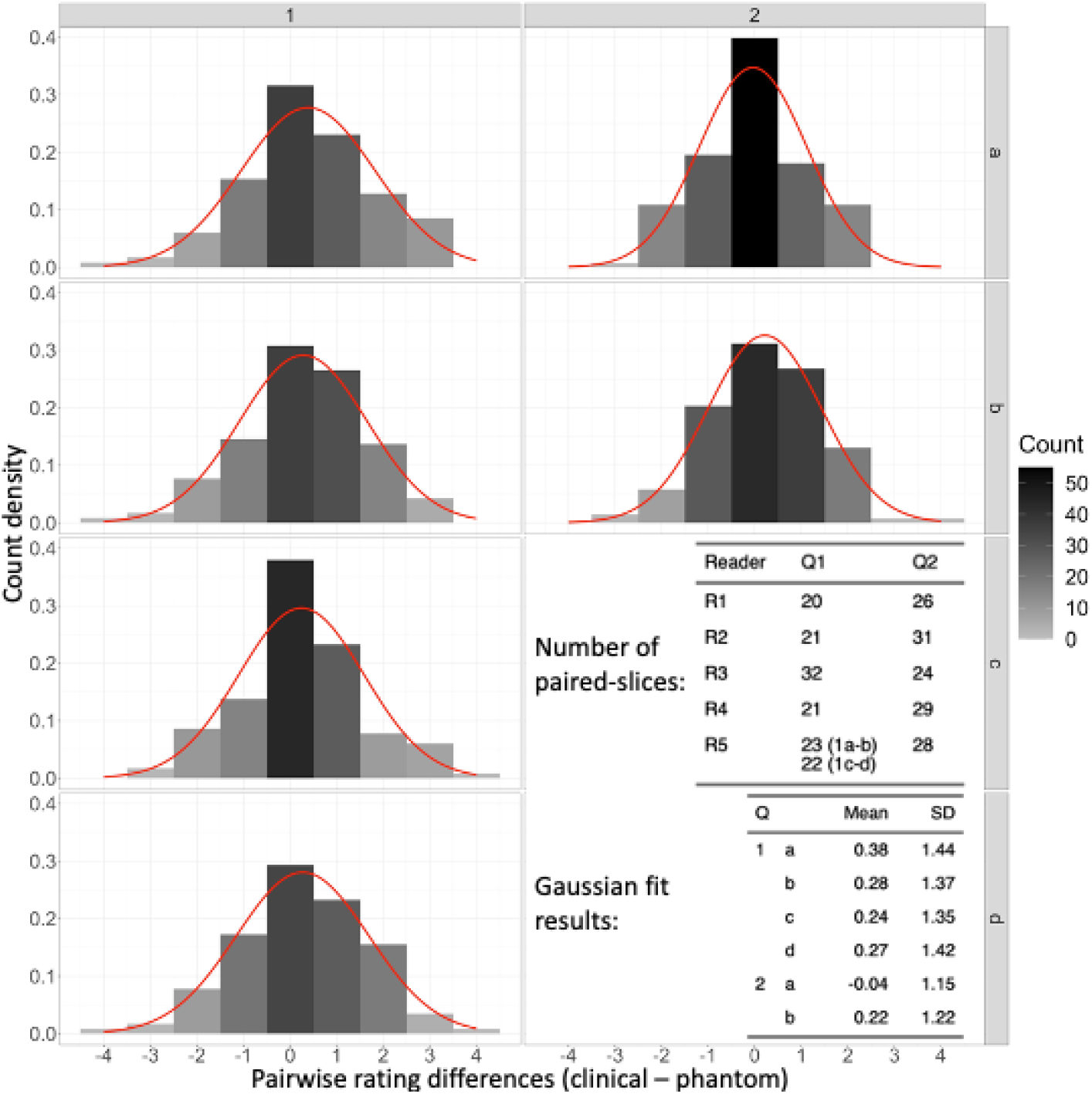
Rating difference frequencies between corresponding (paired) patient and phantom images that were reviewed by the same radiologist, together with gaussian fits to the distributions (red curves). The analysis reveals average differences that are much smaller than a single rating point for all questions and nearly zero points for the COVID-19 severity question (2a).

**Table 2:** Modeling results for mean ratings and mean differences due to having a phantom in the images, rather than a patient, for each of the reader study questions. Results are accompanied by 95% confidence intervals (CI), p-values, and R-squared values.

| Question | Synopsis | Parameter | Estimate | 95% CI | P-value |
| --- | --- | --- | --- | --- | --- |
| 1a | Imaging characteristics | Patient Mean | 3.71 | (3.21, 4.21) | <0.005 |
|  |  | Phantom Eff. | -0.29 | (-0.45, -0.13) |  |
|  |  | R-squared | 0.02 |  |  |
| 1b | Contrast characteristics | Patient Mean | 3.85 | (3.52, 4.19) | <0.005 |
|  |  | Phantom Eff. | -0.27 | (-0.42, -0.11) |  |
|  |  | R-squared | 0.02 |  |  |
| 1c | Noise characteristics | Patient Mean | 3.53 | (3.03, 4.03) | 0.006 |
|  |  | Phantom Eff. | -0.20 | (-0.35, -0.06) |  |
|  |  | R-squared | 0.01 |  |  |
| 1d | Resolution characteristics | Patient Mean | 3.50 | (2.95, 4.05) | 0.006 |
|  |  | Phantom Eff. | -0.22 | (-0.39, -0.06) |  |
|  |  | R-squared | 0.01 |  |  |
| 2a | COVID-19 severity | Patient Mean | 2.77 | (2.49, 3.05) | 0.756 |
|  |  | Phantom Eff. | -0.03 | (-0.2, 0.14) |  |
|  |  | R-squared | 0.00 |  |  |
| 2b | Diagnostic confidence | Patient Mean | 3.56 | (3.1, 4.02) | <0.005 |
|  |  | Phantom Eff. | -0.29 | (-0.47, -0.12) |  |
|  |  | R-squared | 0.02 |  |  |

Assessment of effect sizes with respect to both inter- and intra-reader variabilities are presented in Table 3. The two calculated ratios that were used to estimate the clinical significance of the effect of having a phantom in the image, |*d*′| and |*d*|, are reported for each question separately. For each question, both resulting ratios have similar small magnitudes, with a maximal difference of 0.03, and none surpassing a maximal value of 0.31.

**Table 3:** Assessment of effect sizes with respect to both inter- and intra-reader variabilities, reveal that the effect of having a phantom in the image, rather than a patient, are all smaller than one-third of inter/intra-reader uncertainty, indicating the clinical insignificance of this effect.

| Q | Synopsis | Phantom<br>effect ($\beta_1$) | Inter-reader<br>($\sigma_\varepsilon^2$) | Intra-reader<br>($\sigma_\phi^2$) | $\frac{\vert \beta_1\vert }{\sqrt{\sigma_\varepsilon^2}}$ | $\frac{\vert \beta_1\vert }{\sqrt{\sigma_\varepsilon^2 + \sigma_\phi^2}}$ |
| --- | --- | --- | --- | --- | --- | --- |
| 1a | Imaging<br>characteristics | -0.29 | 0.99 | 0.31 | 0.29 | 0.26 |
| 1b | Contrast<br>characteristics | -0.27 | 0.96 | 0.13 | 0.27 | 0.25 |
| 1c | Noise<br>characteristics | -0.20 | 0.83 | 0.31 | 0.22 | 0.19 |
| 1d | Resolution<br>characteristics | -0.22 | 1.01 | 0.37 | 0.22 | 0.19 |
| 2a | COVID-19<br>severity | -0.03 | 0.83 | 0.09 | 0.03 | 0.03 |
| 2b | Diagnostic<br>confidence | -0.29 | 0.87 | 0.25 | 0.31 | 0.28 |

Results for the reproducibility of PixelPrint’s production process are presented in Figure 5 and Table 4. Figure 5 presents images of two phantoms that were 3D-printed separately using the same patient input (the moderate severity patient), the difference image, and histograms of HU distribution within each image. As can be seen from the figure, differences in HU mainly arise from minor misalignments between the phantoms rather than offsets in attenuation of geometry (Figure 5C). This can also be observed by the excellent overlap of histograms (Figure 5D). Table 4 summarizes SSIM comparisons between the four 3D-printed phantoms. Normalized SSIM values, which were calculated by dividing SSIM values by the ratio of SSIM between the second high-dose scan of phantom #1 and the two other high-dose scans of the same phantom, were between 0.928 and 0.979 with an average of 0.965. This value is higher than the normalized SSIM value of the low-dose scan for phantom #1 (same phantom that was used for normalization).

**Figure 5:**
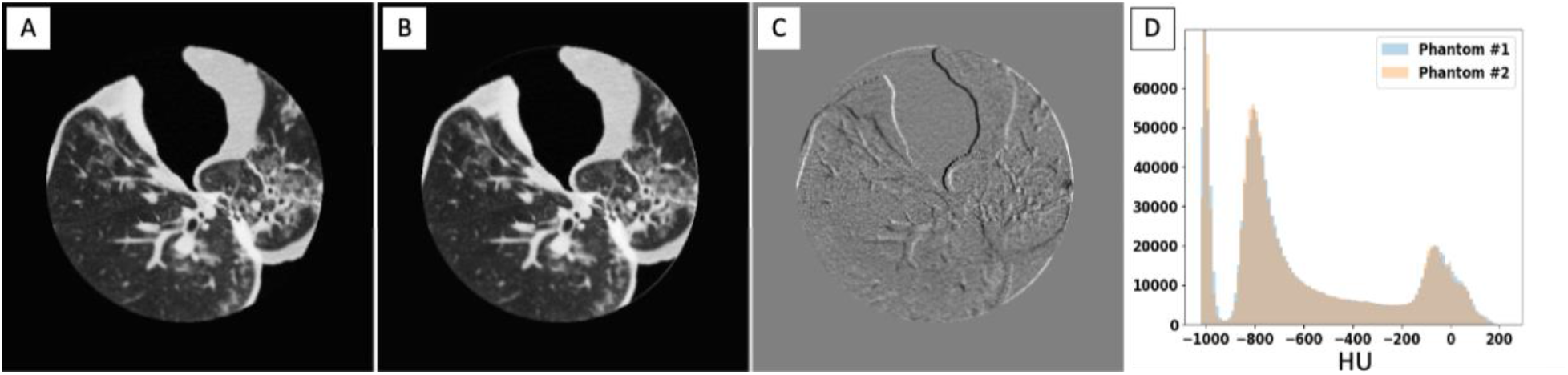
Comparison between two 3D-printed phantoms (A,B), both based on the moderate COVID-19 patient, scanned separately at a high (non-clinical) dose level show high structural similarities and imaging features, implying high reproducibility of the PixelPrint phantom production process. Window level/width are -400/1000 HU. (C) Difference image between the two sets of images reveal that most of the difference between the images are mainly due to slight misalignments between the two phantoms. Window level/width are 0/200 HU. (D) Histograms of HU values within the entire phantom volume demonstrate excellent reproducibility.

**Table 4:**
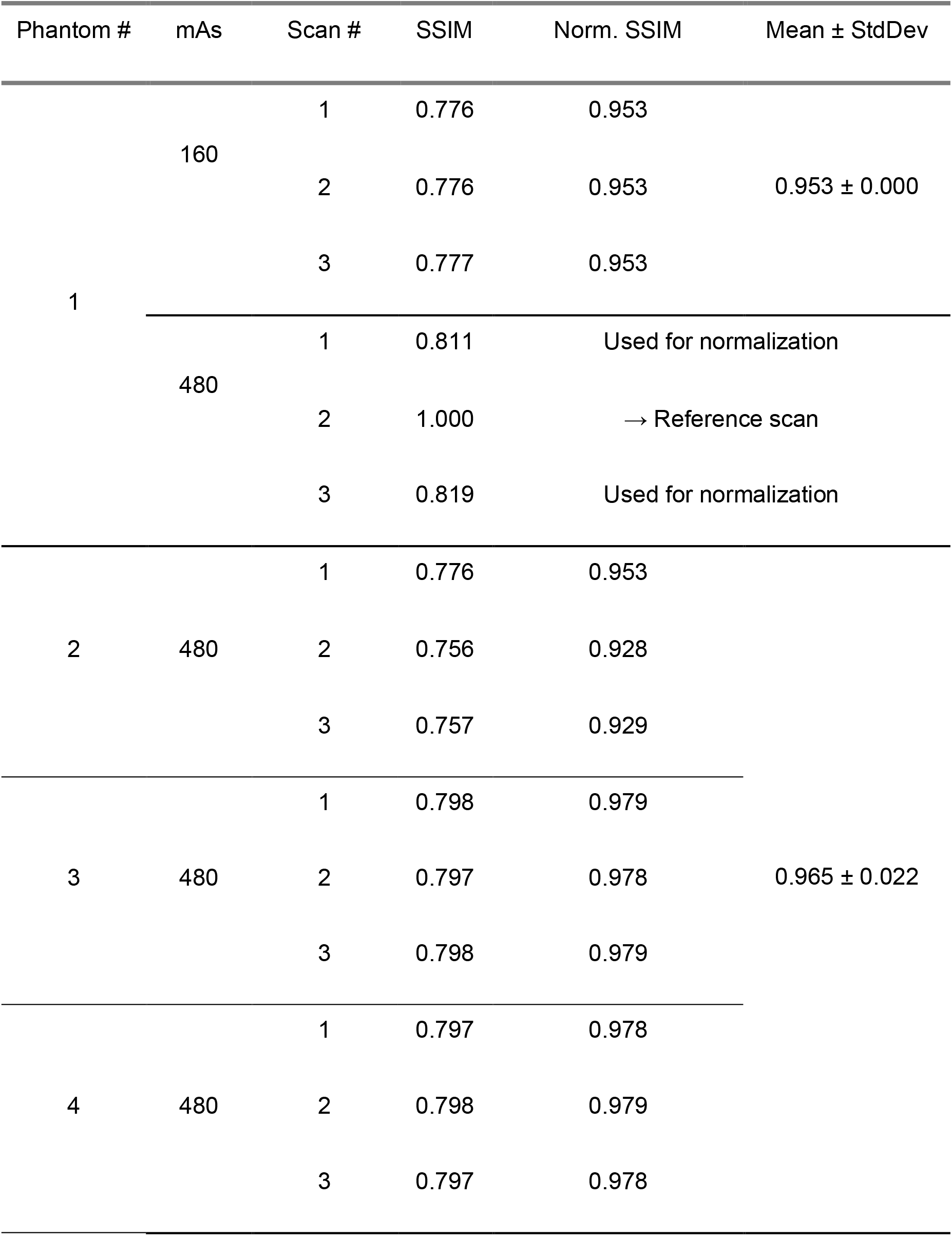
Comparisons of structural similarity index measures (SSIM) between four 3D-printed phantoms that are al based on the same clinical images. Normalized SSIM values, calculated by dividing SSIM values by the ratio of SSIM between the second high-dose scan of phantom #1 and the two other high-dose scans of the same phantom were between 0.928 and 0.979, with an average of 0.965. This value is higher than the normalized SSIM value of the low-dose scan for phantom #1 (same phantom that was used for normalization), demonstrating the high production reliability of PixelPrint.

## DISCUSSION

PixelPrint was developed to provide ground-truth targets for validating the generalizability of inference-based decision-support algorithms between different health centers and imaging protocols, e.g., by imaging the same phantom on multiple scanners, as well as for disease-targeting imaging protocol optimization with realistic patient-based phantoms. We previously assessed the geometrical and attenuation accuracy of our 3D-printed phantoms for CT lung imaging^33^. Here we validated the adequacy of our phantoms for a specific clinical indication, i.e., diagnosis of COVID-19 consolidations, through a “completely blinded” reader study. Statistical analysis of image quality ratings, e.g., imaging characteristics, diagnostic outcome, and diagnostic confidence, revealed that difference in replacing a patient image with a phantom image is, on average, smaller than one-third of a single rating point. Importantly, when examining the clinical significance of these differences by relating them to inter- and intra-reader variability with effect sizes (Table 3), we conclude that the impact of reading a phantom image rather than a patient image is clinically insignificant. Additionally, tests of PixelPrint’s production reproducibility resulted in very high correspondence between phantoms that were 3D-printed using the same patient input. This is based on the higher normalized SSIM values that were measured between high-dose scans of four different phantoms (0.965 ± 0.022), compared to those measured between clinical-dose scans of a single phantom (0.953 ± 0.000).

With many novel modern pattern recognition tools, the improvement in image quality, and the increase in dataset sizes, the field of medical image analysis has grown exponentially in the past decade^4^. Radiomics and clinical decision-supporting AI are emerging as the next frontier for diagnostic and prognostic medical imaging in the new era of precision medicine^2^. The aim of these tools is to automatically extract quantitative information from medical images for assisting evidence-based clinical decision-making^4–6,8^. However, several major challenges hamper the widespread clinical translation of these promising new capabilities. The problem of data variability, which stems from differences in image acquisition and reconstruction settings among medical institutions, and scanner models, is recognized by many as a critical hurdle that requires dedicated solutions to enable the scalability of developed algorithms^6–8^. While recent studies made significant progress with solutions to account for some of the data variability, i.e., normalizations of image quality or imaging features, there is a critical need for lifelike phantoms that will enable the affirmations of these solutions without introducing additional risk to patients or logistical restrictions.

Our study does have limitations. First, while the reader study included a large sample size of images (210 per reader), these images originated from only three clinical patient scans representing three levels of COVID-19 severity. Second, our study focused on a specific clinical indication, i.e., diagnosis of COVID-19 pneumonia. Further studies are required to validate the adequacy of PixelPrint for other lung imaging indications, e.g., lung nodule detection. Nevertheless, our results provide compelling evidence that PixelPrint can readily serve as an accurate tool for optimization of disease-targeting protocols and for experimental validation of novel inference algorithms, such as radiomics and predictive AI.

In conclusion, we have demonstrated PixelPrint’s ability to produce realistic 3D-printed phantoms reliably. As the utilization of these phantoms will grow, they will become more beneficial to the entire community and enable standardization of tests and comparisons of evaluation of advanced medical inference algorithms. For this, we offer copies of the phantoms presented in this study, as well as phantoms based on specific CT images, for the larger medical, academic, and industrial CT community (visit www.pennmedicine.org/CTResearch/PixelPrint).

## Data Availability

All data produced in the present study are available upon reasonable request to the authors

## ACKNOWLEDGEMENT

We acknowledge support through the National Institutes of Health (R01-CA-249538, R01-CA-264835-01, and R01-EB-030494).

